# Time to recovery of asphyxiated neonates and it’s predictors among newborns admitted to neonatal intensive care unit at Debre Berhan Comprehensive Specialized Hospital, Ethiopia

**DOI:** 10.1101/2024.01.28.24301901

**Authors:** Sisay Girma Yehouala, Esubalew Tesfahun, Tadesse Mamo Dejene, Zenebe Abebe Gebreegziabher

## Abstract

**Background:** Even though there have been inquiries into the survival rates of asphyxiated neonates in Africa, there is scarce data concerning the recovery duration for asphyxiated newborns in developing nations and the factors affecting this process. Consequently, the objective of this study is to ascertain the time it takes for asphyxiated neonates to recover and identify its predictors.

**Methods:** Conducting a retrospective follow-up investigation, the study took place at Debre Berhan Comprehensive Specialized Hospital from January 1^st^, 2020 to December 31^st^ 2022, involving a sample size of 330. The analysis included the computation of the Kaplan-Meier survival curve, the log-rank test, and the median time. Additionally, a multivariable Cox proportional hazard regression model was employed to determine the survival status.

**Results:** in this study, among the 330 participants (100%), a total of 270(81.8%) successfully survived throughout the entire cohort. Predictors are independent of each other, affecting the time to recovery and survival of asphyxiated neonates, encompassed prolonged labor (AHR: 0.42, 95%CI:0.21-0.81), normal birth weight (AHR:2.21, 95% CI: 1.30-3.70),non-altered consciousness (AHR:2.52, CI:1.50-4.24), non-depressed moro reflex of the newborn (AHR:2.40, 95%CI: 1.03-5.61), stage I HIE (AHR: 5.11, 95% CI: 1.98-13.19), and direct oxygen administration via the nose (AHR: 4.18, 95% CI: 2.21-7.89).

**Conclusion:** The duration for recovery seems to be slightly prolonged in comparison to other research findings. This underscores the significance of vigilant monitoring, early preventive interventions, and swift actions to avert the progression of infants to the most severe stage of HIE.

## Introduction

Perinatal asphyxia is the inability to initiate and maintain breathing at birth, characterized by significant impairment of respiratory gas exchange (oxygen and carbon dioxide), resulting in progressive hypoxemia and hypercapnia, as well as significant metabolic acidosis. A newborn with a fifth minute Apgar score of seven is diagnosed with birth asphyxia. According to the World Health Organization (WHO), birth asphyxia can be defined and classified as mild, moderate, or severe using the APGAR score at 1 and 5 minutes. During the 72 hours period following the hypoxic-ischemic insult, newborns may develop mild, moderate, or severe encephalopathy [1–4].

The global incidence of perinatal asphyxia is higher in developing countries than developed countries[5]. Perinatal asphyxia can be caused by maternal or fetal conditions that occur before, during, or after birth, or by a combination of these[6, 7]. Perinatal asphyxia causes a variety of outcomes in the neonate’s life, including multi-organ dysfunction, death, severe neurodevelopmental delay, motor delay, cerebral delay, and hypoxic-ischemic encephalopathy (HIE)[8, 9].

The magnitude of asphyxiation at birth is ten times more frequent in developing countries compared to developed countries[1]. Based on the WHO estimates, 3.6 million infants (3%) have moderate to severe asphyxiation at birth, with 23% dying and about the same number suffering from severe sequelae in developing countries [10]. In the western world, there is a higher likelihood of survival for asphyxiated neonates through improved perinatal care, ongoing fetal monitoring during labor, early presentation of newborns to Neonatal Intensive Care Units (NICUs), and treatment options, there is higher likelihood of survival for asphyxiated neonates. This was not true in developing nations, where mortality rates are high and recovery times or average hospital stays are prolonged[11]. The majority of neonatal deaths are significantly caused by perinatal asphyxia (PNA). Similar to this, Ethiopia continues to have a high burden of birth asphyxia (22.52%), which has been identified as the second leading cause of neonatal mortality [12].The study done in a hospital of Southeast Nigeria found that 61.3% of newborns survived and were sent home [13]. Another retrospective study done in Nigeria showed that 25.5% of newborns died and 63.9% of newborns were discharged[14]. The prospective case-control study in Cameroon with 90 cases of asphyxia reported that 81.1% of the affected infants had successful outcomes, 12.2% were discharged with complications, and 6.7% died during the study period [15]. According to a study conducted in Ghana, of the 289 asphyxiated infants admitted to the Korle-Bu Teaching Hospital NICU during the study period, 78.2% survived, and the other infants perished[16]. Despite this fact, there is limited evidence about the survival of neonates having asphyxia and associated risk factors that affect the survival of neonates with asphyxia. The purpose of this study was to determine the survival of neonates having asphyxia and its predictors. Identifying predictors that affect the survival status of neonates has been recognized as the most important preventable and manageable cause of neonatal mortality. The identification of asphyxiated newborns at birth could help implement interventional projects, develop preventive measures and treatment protocols to lessen the effects of its complications, as well as significantly reduce the length of hospitalization and mortality linked to this condition.

## Methods

### Study area and settings

The study was carried out at Debre Berhan Comprehensive Specialized Hospital, Ethiopia. The hospital has a tertiary-level neonatal intensive care unit and serves over 3 million people with preventive, curative, and rehabilitative services. It also provides delivery service 24 hours a day, 7 days a week, and in 2022 there were 4277 deliveries annually. In 2022, the NICU ward’s annual perinatal asphyxia (PNA) admission was 214, and there are 28 beds, 5 incubators, and 15 radiant warmers with 15 comprehensive BSc nurses, 13 neonatal nurses, 3 pediatricians, and 4 general practitioners.

### Study area and period

A retrospective cohort study was conducted on asphyxiated newborns admitted to Debre Berhan Comprehensive Specialized Hospital (DBCSH), neonatal intensive care units (NICUs). The study was conducted from April to May 2023 and used data from the registration log book and patient charts.

### Sample size determination and Sampling technique

The sample size would be determined by the log-rank test, and Freedman method using STATAsoftwareversion14.0.

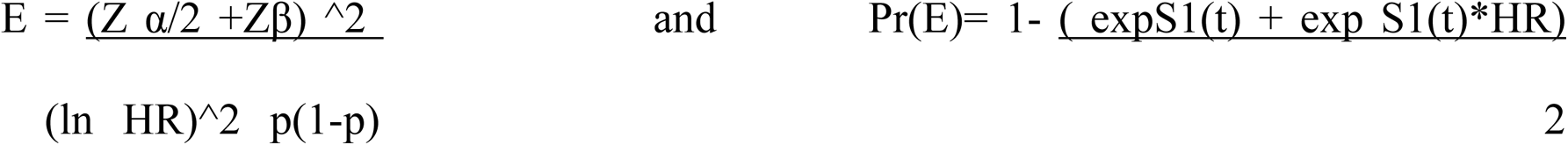

Where:

Zα/2 = standard normal variable at 95% confidence interval level = (1.96) Zβ = power of 80% =(0.842), P= cumulative survival probability at end of study (28 days) is 0.04%, Hazard ratio= 0.5, Pr (E)= probability of event for outcome variable(recovery) and IR = incidence rate is 10% as obtained from a retrospective study done in Addis Ababa public hospitals on time to recovery and predictors of asphyxia among neonates[17]. E = number of interested events is 66 and the probability of an event (recovery) is 0.2 so the total sample size is calculated as;

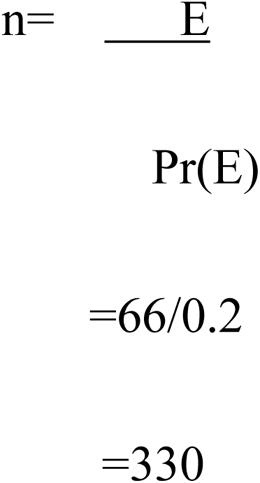

The samples were taken in a way that the registration logbook was searched for medical record numbers of babies diagnosed with perinatal asphyxia. The study participants were then choosen using a computer-generated simple random sampling technique from the isolated medical record numbers in the hospital. Finally, from April to May 2023, the selected medical charts would be reviewed

### Data Collection

A structured data extraction checklist/sheet was developed using the Ethiopia Ministry of health standardized log, registration book, perinatal follow-up chart, neonatal evaluation sheet, neonatal referral paper, and other reviewed articles[17]. The participants in the study would then be selected based on eligibility criteria, and all available information from patient records was reviewed. The pre-test, which would be performed on a population equivalent to 5% (17 charts) of the study sample, was ensure data quality. Errors would be discovered during the verification process and were corrected and modified before the analysis to ensure agreement of the data abstraction format and the research objectives during data management, storage, cleaning, and analysis.

### Data Processing and Analysis

The collected data were coded, edited, cleaned, and entered on a Google spreadsheet and finally exported to the statistical software STATA version 14.0 for further analysis. The data would be presented using descriptive and inferential statistics. Schoenfeld’s residual test would be used to validate the Cox-proportion hazard regression models’ necessary assumptions. To estimate survival time, the Kaplan-Meier survival curve would be used, and the Log-rank test would be used to compare survival curves between categorical predictors. A bivariate Cox-proportional hazards regression model would be fitted for each explanatory variable and variables with p-values of < 0.2 were then fitted to a multivariable Cox-regression analysis to identify independent predictors of asphyxiated neonate survival. The statistical significance of predictor associations was determined using a hazard ratio with a 95% confidence level. In the multivariable cox-regression analysis, variables with a p-value of < 0.05 were considered significant predictors of asphyxiating neonate survival.

## Results

### Socio-demographic and Obstetrics characteristics of the study participants

Three hundred thirty charts of asphyxiated neonates were reviewed; 330(100%) were eligible for this study. The majority 238 (72%) of the mothers were between the ages of 20 to 34. More than half of the mothers 205(62%) gave birth for the first time, and nearly all of the 319(97%) pregnant women had antenatal care visits. Regarding the method of delivery, 204 (62%) and 54 (16%) were spontaneous vaginal delivery and cesarean section (C-section) respectively. Pregnant women who had obstetrics and medical complications recorded during their pregnancies had pre/eclampsia in 211 (64%) cases, meconium-stained amniotic fluid (MSAF) in 87 (26%) cases, oligohydramnios in 19 (6% cases), and a history of abortion in 13 (4% cases). Of the total 330 newborns admitted, 60% of them were born in Debre Berhan compressive specialized hospital (inborn) (Table 1).

**Table 1:**
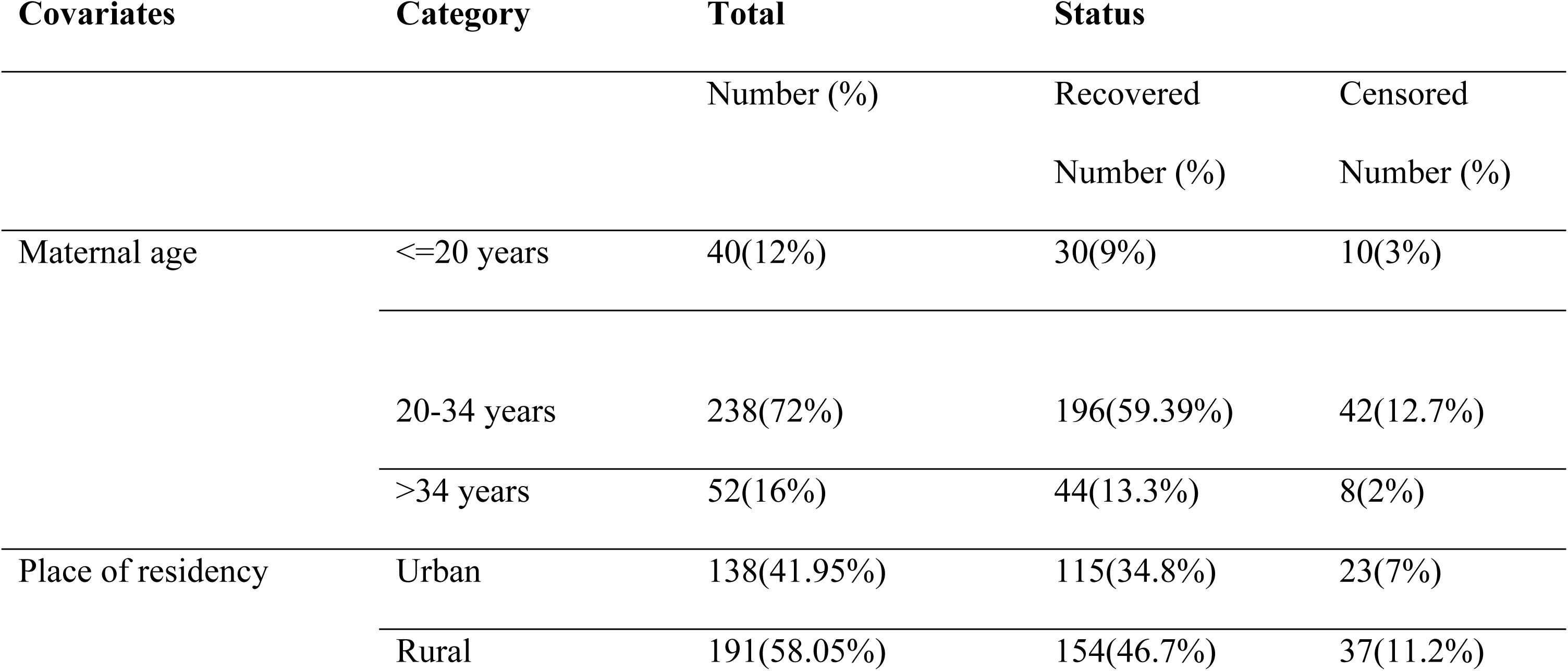
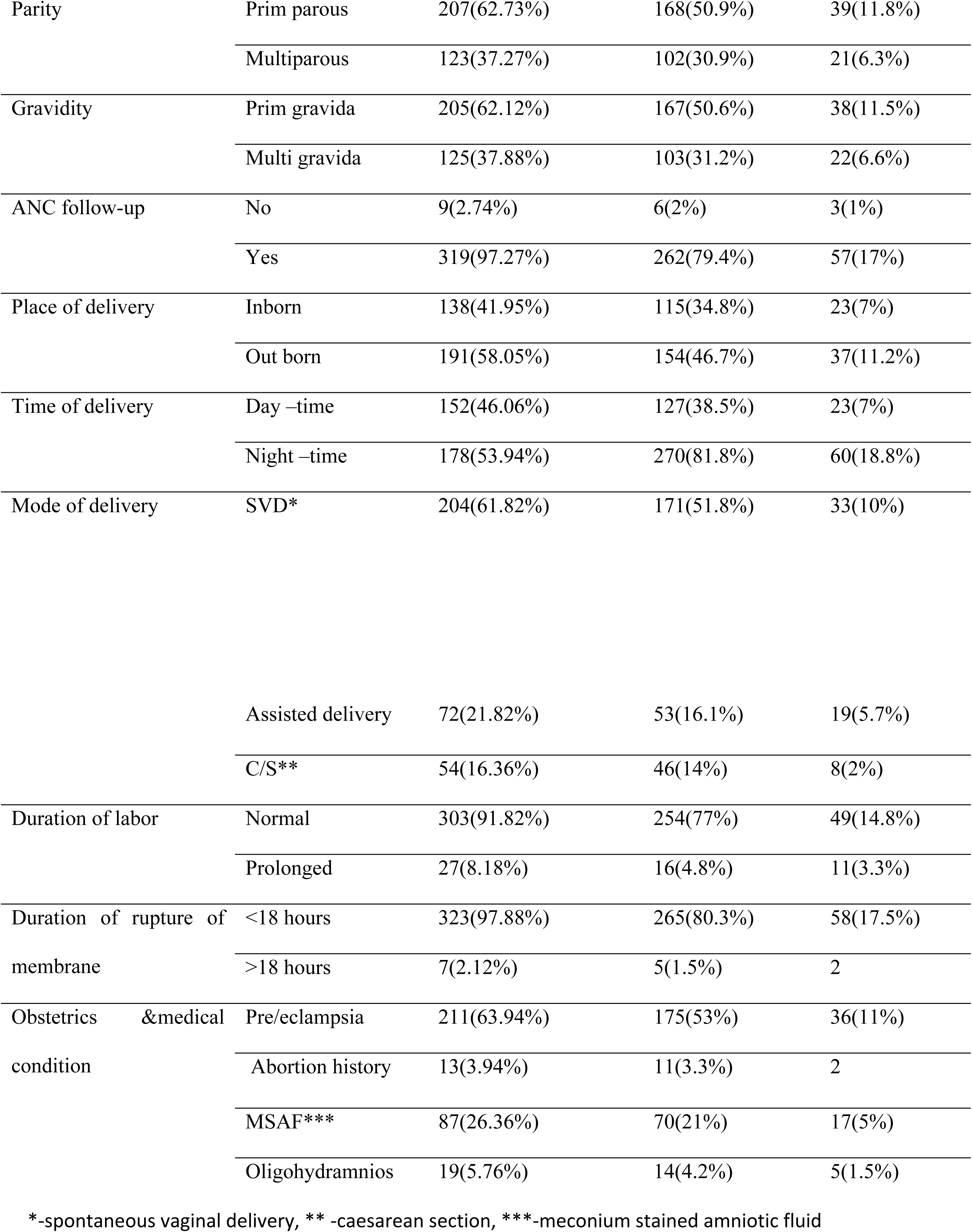
Socio-demographic and obstetrics characteristics of the study participants, Debre Berhan Comprehensive Specialized Hospital (DBCSH), 2020-2022 (n=330), 2023.

### Neonatal characteristics of the study participants

In the cohort, 134(60%) of the participants were male babies. From the cohort 44(13%), 264 (80%), and 22(7%) were preterm, term and post-term respectively. According to their postnatal age, those who presented before 24 hours in the neonatal ward were 268(81.21%), and more than 24 hours were 62(18.79%). Nearly three-fourths of the neonates didn’t cry immediately at birth and one-fourth of neonates cried and had normal APGAR scores of (>=7) at the first minute of their life in both survival and censored. Regarding the 5^th^-minute an APGAR score, 218 (66.06%) had APGAR score <7 and 112(33.94%) of them had APGAR score of >=7. From the cohort, in the case of their birth weight 66(20%) were low birth, and 264(80%) of them had normal birth weight (Table 2).

**Table 2:**
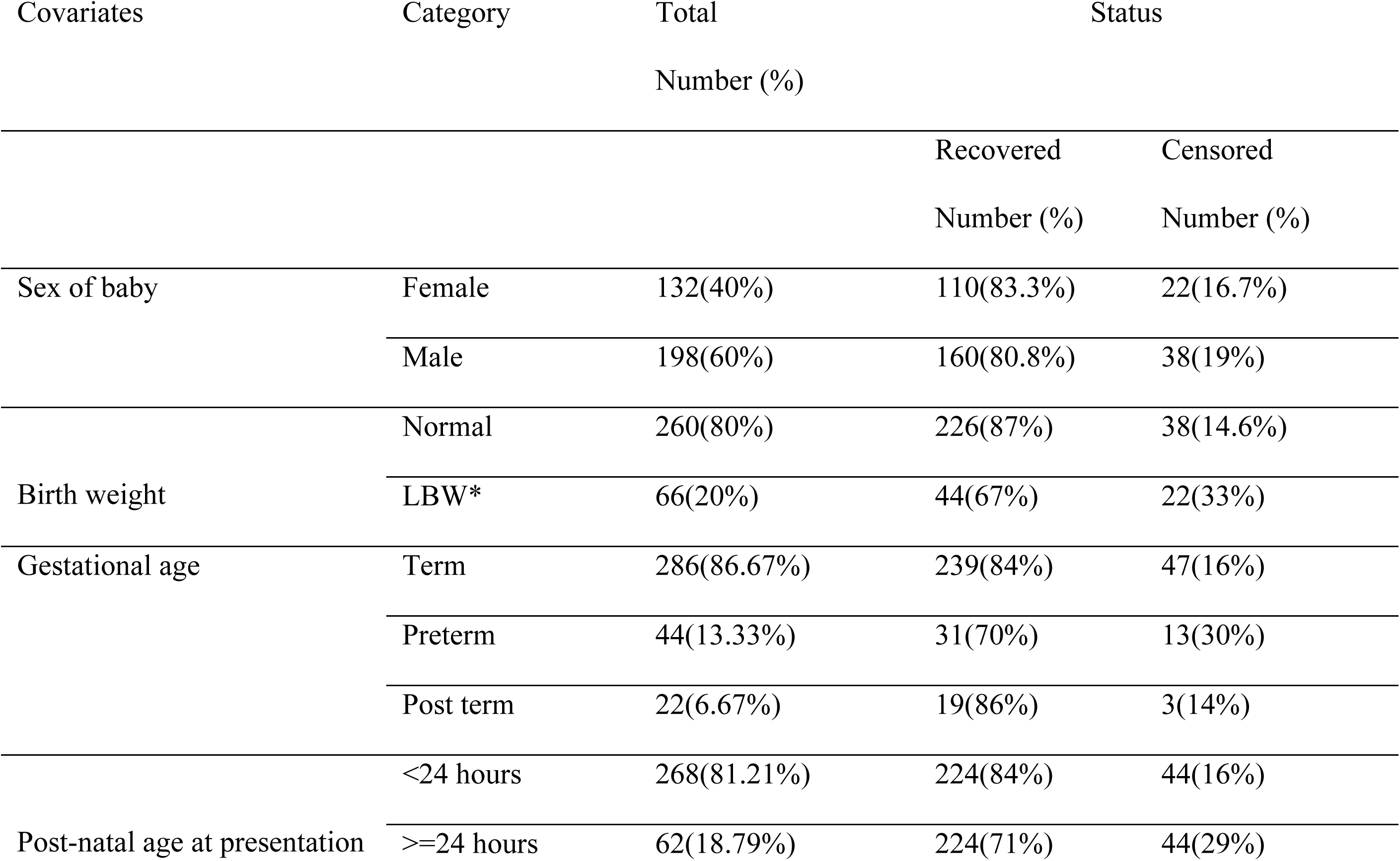
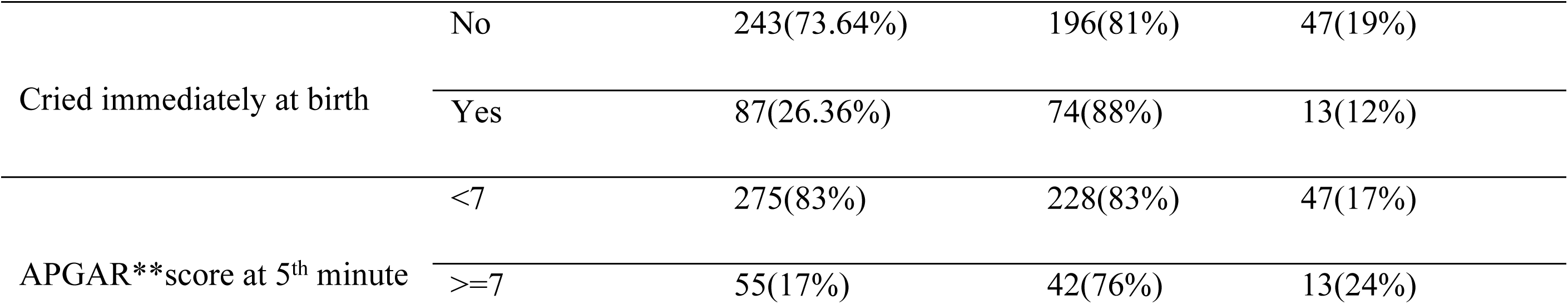
Neonatal characteristics of the asphyxiated babies who were admitted to the NICU of DBCSH, Ethiopia, 2020-2022 (n=330), 2023.

### Clinical and Laboratory characteristics of the study participants

Respiratory distress syndrome 198(60%), hypothermia 82(24.85) and meconium-stained amniotic fluid 50(15.15%) was the most frequently observed additional medical issues among asphyxiated newborns at admission and throughout their hospital stays. 36(15%) of the newborns with stage II HIE were censored. 210 (64%) and 60 (18%) of the total neonates who survived were still alive when they were discharged from stage II and stage I, respectively. Of the neonates who survived showed up in NICUs with altered consciousness 77(23.33%) and 54(20%) had a depressed moro reflex of them were censored. At admission 59(17.88%) had gasping type of breathing or respiratory rate less than 30 breathe per minute, 218(66.06%) were hypothermic and 153(46.36%) had low oxygen saturation respectively. Out of the total survived neonates 104(38.5%) had normal respiratory rate, 170(52%) had normal heart rate, 80(24.2%) had normal temperatures and 143(43%) had normal oxygen saturation levels within 24 hours of postnatal age. After 24 hours of admission 8(2.42%), 44(13.33%) had had respiration rate <30b/min and >60b/min respectively. Among those whose heartbeats were >160 b/min were 18(5.45%) and those 124(37.58%) were hypothermic (<36.5oc), and 25(7.58%) were desaturated within the room air or saturation level <85%.

### Survival status of asphyxiated neonates

A total of 2706 neonates-days, ranging from 1 day to 18 days, were spent monitoring the health of 330 admitted asphyxiated infants. The median survival time was 9 days (95% CI: 0.82-0.93). From the total, 270 (81.8%) of the asphyxiated neonates in the cohort survived, and 60 (18.2%) were censored. According to this study’s overall survival rate (incidence density), which was 9.9 per 100 neonates-days of observation (95% CI: 8.85-11.24), the estimated cumulative survival at 1, 3, 7, 14, and 18 days was, respectively, 1.0, 1.0, 0.69, 0.03, and 0.0.

### Predictors of Survival status of asphyxiated neonates

The results of a multivariable Cox regression analysis showed that women who had babies from prolonged labour were 58% less likely to recover from asphyxia than their counterparts (AHR: 0.42, 95% CI: 0.21-0.81). Babies with normal birth weight were 2.2 times more likely to recover from asphyxia than those with low birth weight (AHR: 2.21, 95% CI: 1.30-3.76). Newborns who hadn’t altered their level of consciousness were 2.5 times more likely to recover faster from asphyxia than their counterparts (AHR:2.52, 95%CI: 1.50-4.24) and those who did not face depressed moro reflexes were 2.4 times faster to recover from asphyxia than the counterparts (AHR: 2.4, 95%CI:1.03-5.61). As the HIE stage increased, the recovery process slowed or got longer. Newborns with stage I HIE were 5.1 times more likely to recover faster from asphyxia than their counterparts (AHR: 5.11, 95% CI: 1.98-13.19). According to a resuscitation and management protocol aminophylline is one option given for newborns and those who took aminophylline were 2 times more likely to recover than their counterparts (AHR: 2.05, 95% CI: 1.06-3.96). Newborns who had received direct oxygen (intranasal) 4.2 times more likely to recover faster from asphyxia than those who had continuous positive airway pressure (CPAP) (AHR: 4.18, 95 CI: 2.21-7.89). Vital signs were a key indicator of whether a newborn was recovering from asphyxia or deteriorating, and the results of the multivariable Cox regression analysis showed that asphyxiated babies’ respiratory rates at 24 hours (30-60 breath/min) were 8.6 times more likely to recover faster from asphyxia than the counterparts (AHR:8.68, 95%CI: 3.78-19.90). Babies with heart rates <100 beats/minute at 24 hours were 82% less likely to recover earlier than their counterparts (AHR: 0.18, 95% CI: 0.37-0.93) and babies with heart beats between >160beat/minute were 79% less likely to recover from asphyxia than their counterparts (AHR: 0.21, 95% CI: 0.08-0.50). As asphyxia is a deprivation of oxygen to the brain measuring the level of saturation was mandatory. From the multivariable those babies who had oxygen saturation level <85% at admission were 44% less likely to recover earlier from asphyxia than their counterparts. Regarding comorbidities, neonates who had random blood sugar (RBS) (>60 mg/dl) at 24 hours had 42-fold more likely to recover faster from asphyxia than their counterparts (AHR: 6.2, 95%CI: 3.54-15.56) (Table 3).

**Table 3:**
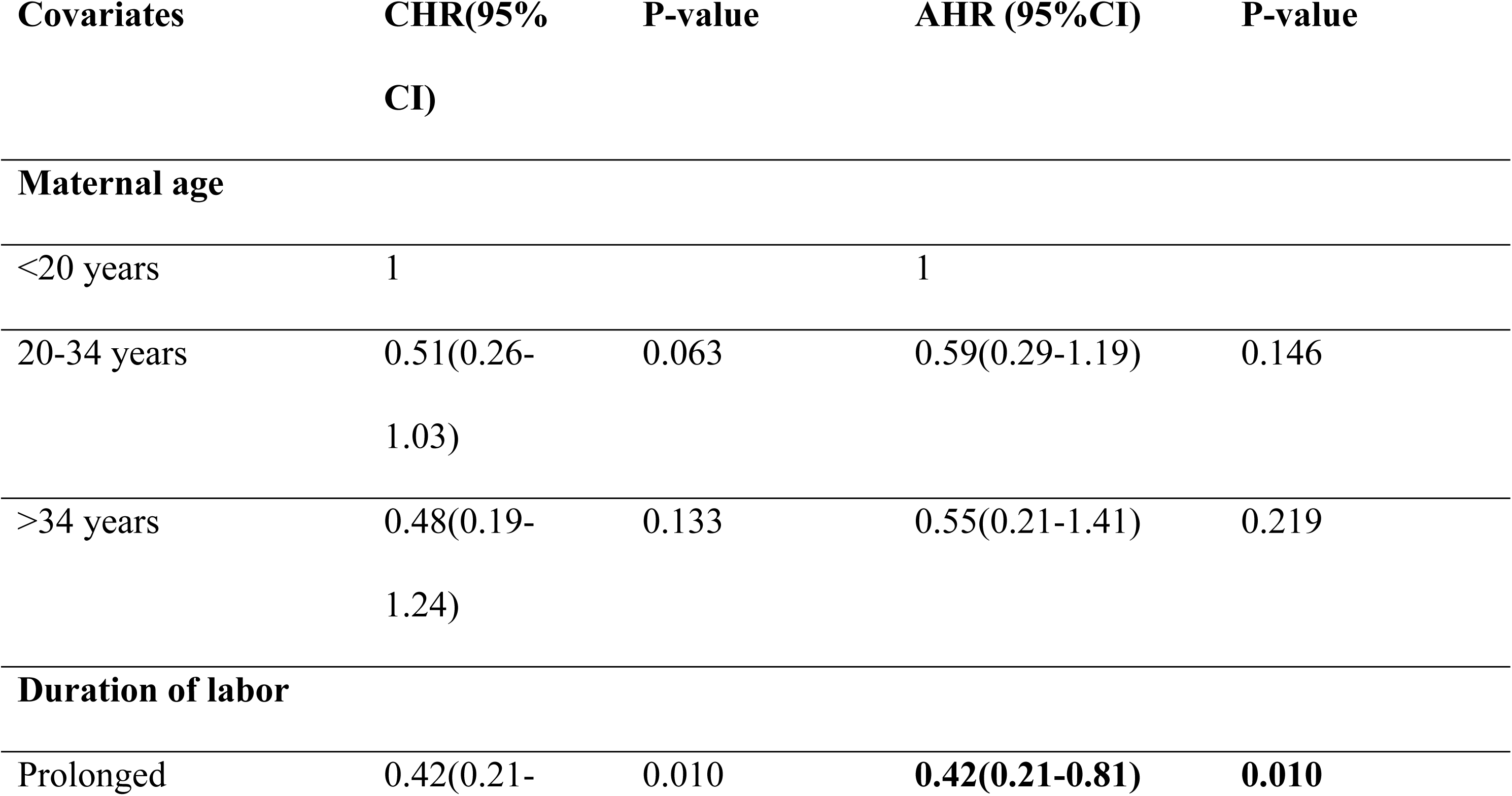
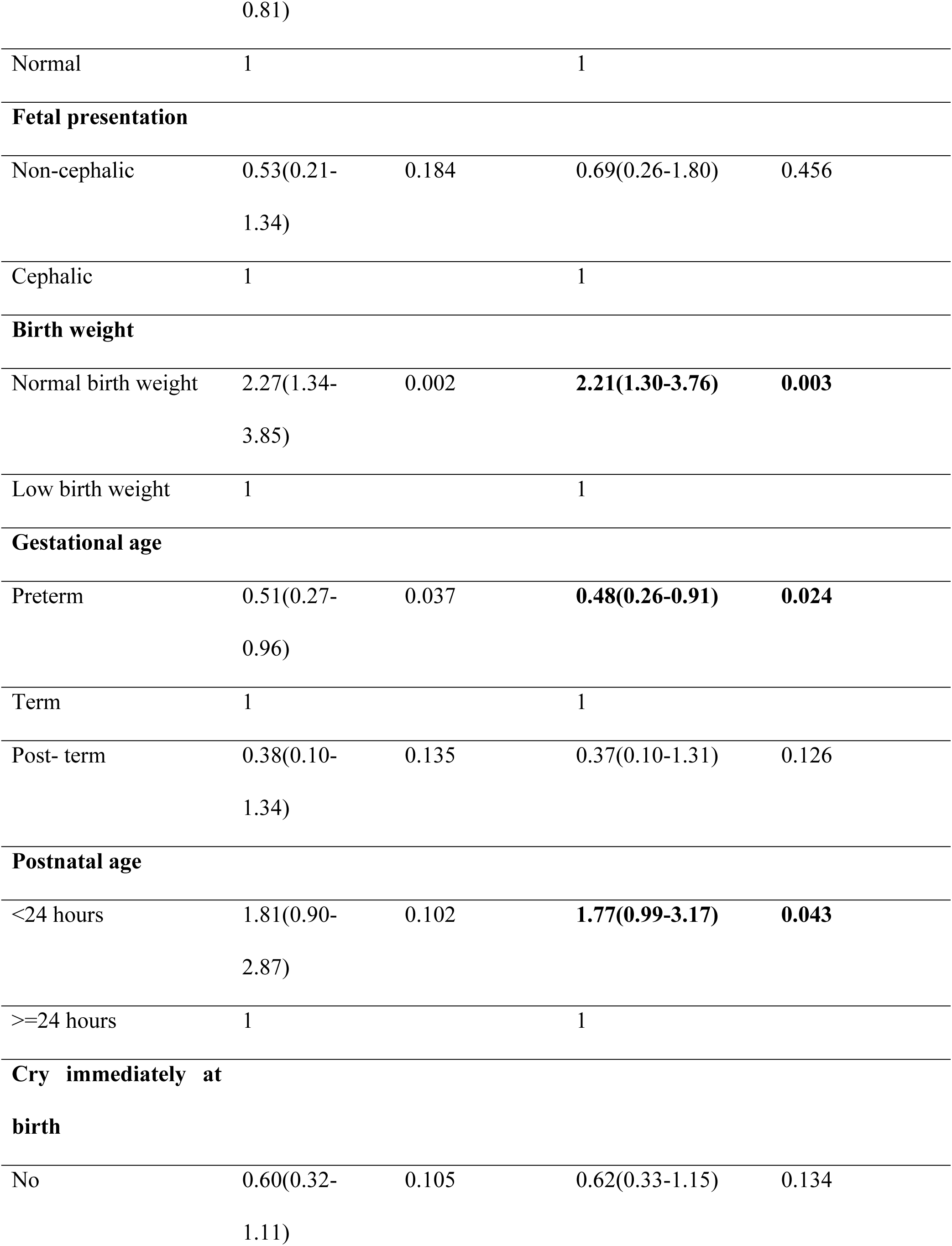
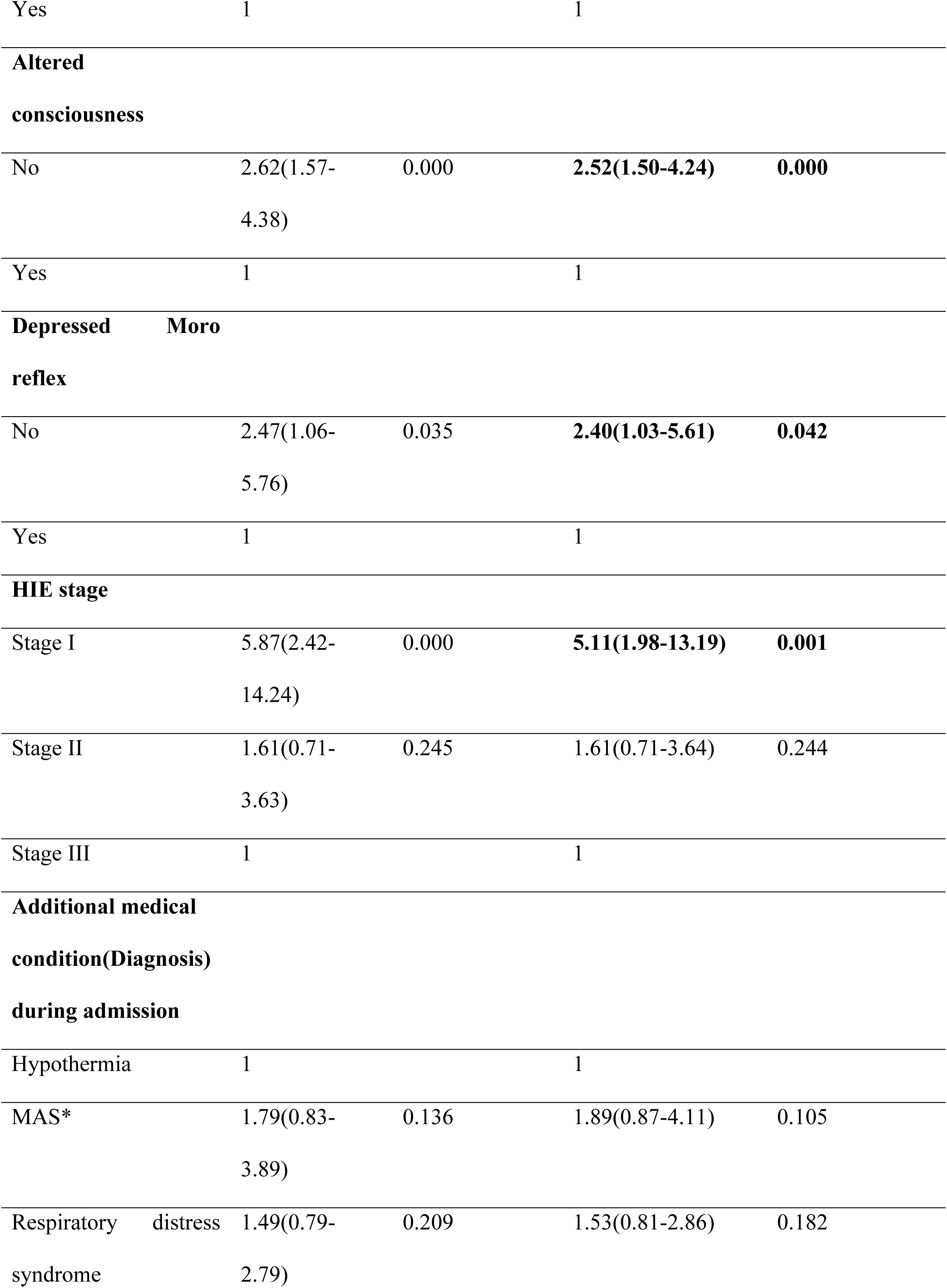
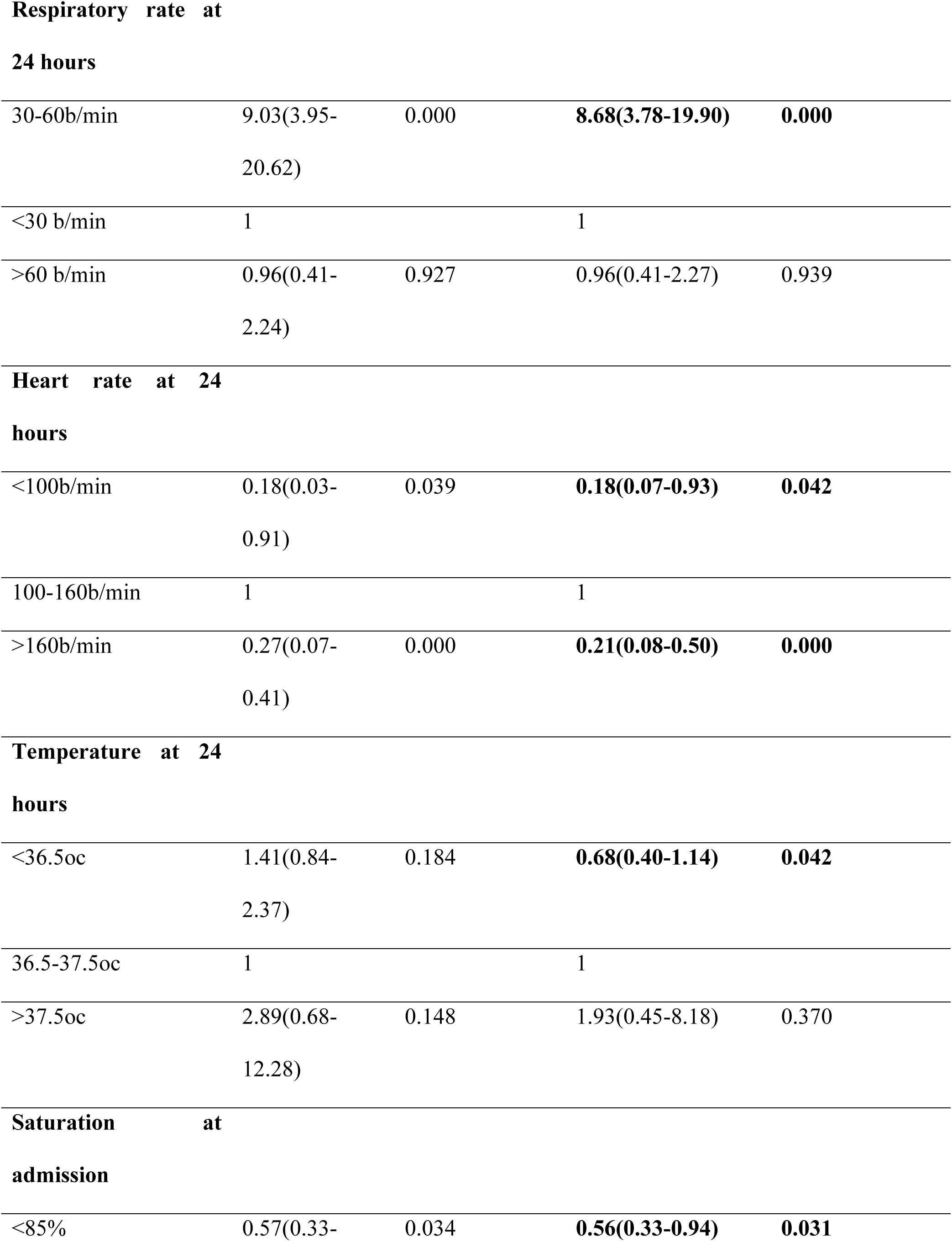
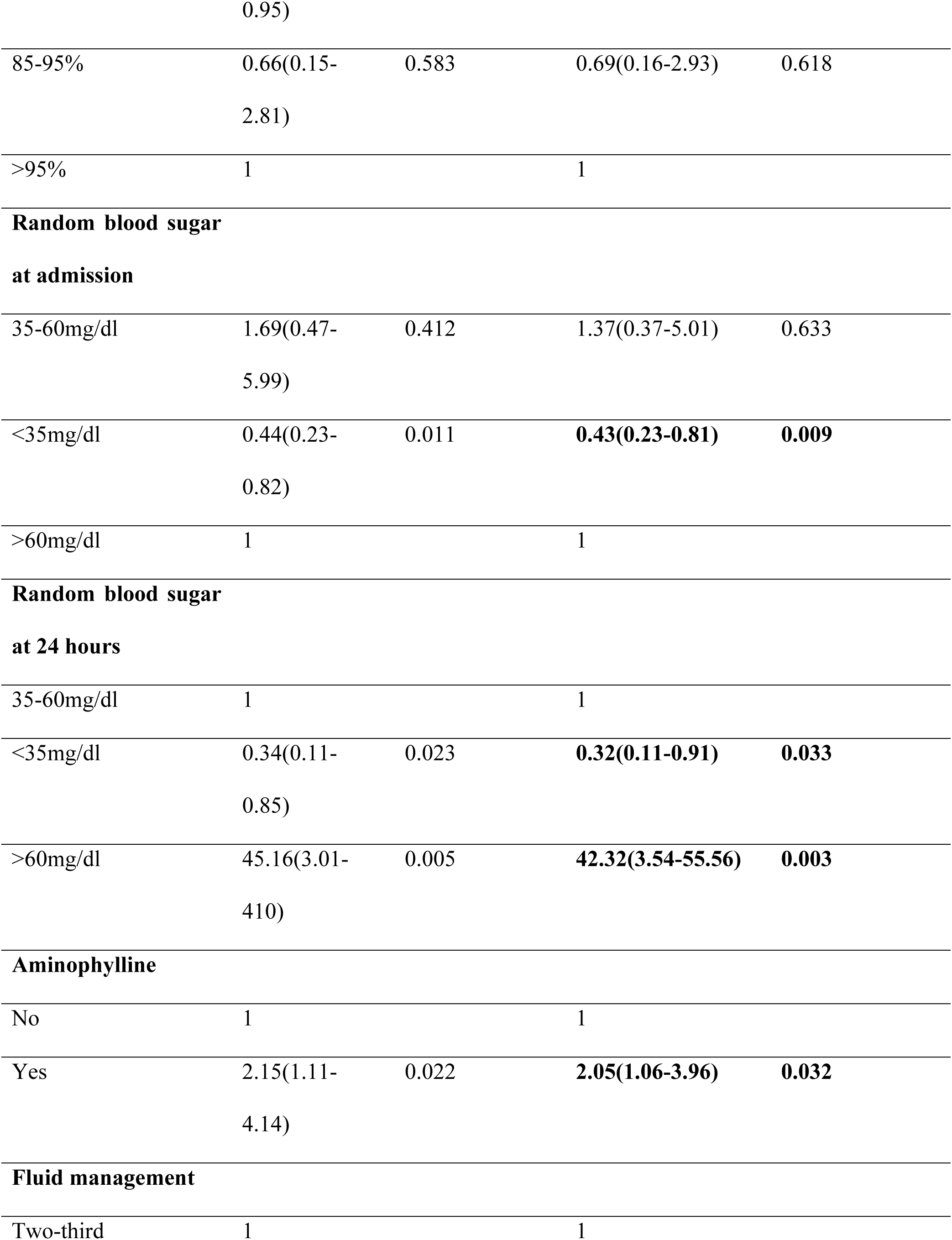
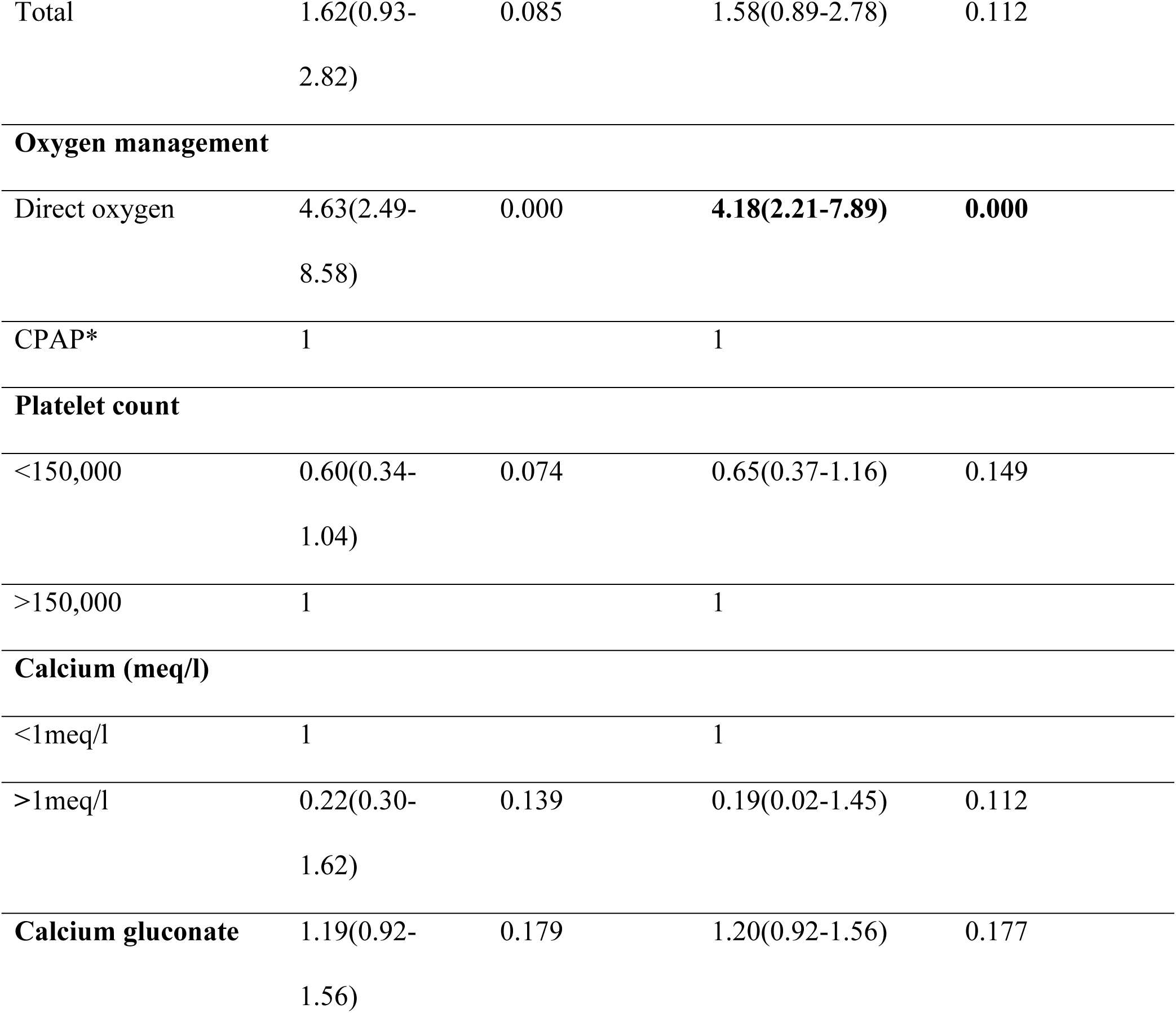
Bi-variable and Multivariable Cox regression analysis results of asphyxiated babies who were admitted at NICU of DBCSH, Ethiopia, 2020-2022 (n=330), 2023.

## Discussion

The current study presented the time to recovery from asphyxiated neonates and its predictors among asphyxiated neonates admitted to Debre Berhan Comprehensive Specialized Hospital. This study found that the median recovery time from asphyxiated newborns was 9 days. This finding was comparable with the study in Addis Ababa (8 days) [17], and Dire Dawa (7 days) [18]. This study showed that preterm and low birth weight neonates have a lower chance of surviving compared to their counterparts, and the result is statistically significant.

The scientific and clinical explanation for this is that low birth weight and/or preterm babies typically have feeding issues, immature thermoregulation, and respiratory center, and poor immune systems, and not providing standardized NICU care may lead to a poor prognosis and a lengthened hospital stay. The gestational age, birth weight, and age of the neonates at the time of presentation to the NICU had an impact on the asphyxiated newborns’ survival rates and recovery times, which is related to the prematurity of the respiratory center, nervous system, immune, and thermoregulation mechanism. Findings were comparable to the study conducted at Jimma and Kenya[19, 20]. The results of this study showed that, depending on the stage of hypoxic-ischemic encephalopathy (HIE), the recovery and survival rates of newborns who had been asphyxiated varied. There is a marginally higher chance of surviving and recovering from asphyxia in stages I HIE than the counterparts. This conclusion was supported by additional findings [21–25]. The scientific explanation for this finding could be, primarily, in fact, asphyxia is followed by the renal system and the central nervous system as the most severely affected systems. The primary and secondary energy failure that frequently occurs as HIE progresses reduces the prognosis of asphyxiated infants and causes the emergence of various neurologic sequelae[26]. Second, management and care are frequently inadequate and ineffective when the neonate has multiple organ damage and prolonged hypoxia. Finally, infants with advanced HIE were frequently kept on NPO (nothing by mouth) and had unstable blood sugar levels out of concern for necrotizing enterocolitis, which aggravated malnutrition and thrombocytopenia. This might complicate the problem, necessitating more time and resources to address any resulting issues. This study’s findings also showed that babies’ vital signs change during admission and at 24 hours duration of admission (respiratory rate at 24 hours of admission, heart rate at 24 hours, temperature at 24 hours, saturation at admission, random blood sugar at admission, random blood sugar at 24 hours) statistically significant. This vital sign change results from when babies haven’t got enough oxygen during birth and end up with permanent injury to the brain, heart, lung, kidney, bowel, and other organs. So, from this finding, if babies hadn’t gotten management earlier, the recovery time of the babies would have declined. Similar findings were studied in Nigeria[27].

Statistics from this study revealed that the patient’s clinical condition upon admission—an altered level of consciousness and a depressed moro reflex was statistically significant and had an impact on the recovery of the patient. South Africa and Nigeria[14, 25], two resource - constrained nations, also supported the findings. And also, asphyxiated newborns who received direct oxygen through the nose (AHR= 4.18, 95 CI: 2.21-7.89) and aminophylline (AHR=2.05, 95 CI: 1.06-3.96), which enhances the recovery of babies from asphyxia than that of their counterparts. This finding was contradicted by studies conducted in Tanzania[28, 29]. This can be scientifically explained if the babies are supported with intranasal oxygen means that the insult of the organ is mild and can be reversed easily and it helps to maintain a normal state. Whereas if babies received CPAP (continuous positive airway pressure), the insult is severe and might be difficult to reverse. According to this study prolonged duration of labor is a significant factor for recovery of asphyxia (AHR: 0.42, 95% CI: 0.2-0.81). As the duration of labor was prolonged the chance of babies being asphyxiated was higher and the likelihood of recovery from asphyxia decreased. This finding is supported by a meta-analysis study in Ethiopia[30]. And explained by excessive contraction leads to meconium leaks and the neonate may suck and the umbilical cord might be compressed and either ischemic or bleed. Finally, end up with asphyxia. The administration of calcium gluconate during the first few days of life along with intravenous fluid therapy may have potential benefits for the baby and improve survival chances compared to those who do not take prophylactic calcium gluconate, even though the current study’s statistical findings are not statistically significant in this regard. The most common scientific causes of hypocalcemia in newborns with asphyxia are low calcium intake, hyperphosphatemia, too much bicarbonate, and functional hypoparathyroidism[31]. In a Tanzanian randomized control trial to assess the effectiveness of prophylactic calcium gluconate administration in the first five days of life, the need for hypocalcemia management was reduced in newborns who received intravenous calcium infusions[32].

## Conclusion

In this finding, more than three-fourths of asphyxiated babies had positive outcomes (recovered) during the entire cohort. Besides, the overall incidence rate of recovery was 9.9 per 100 neonates-days observations, Prolonged labor, low birth weight, gestational age, postnatal age at presentation, altered consciousness at admission, depressed Moro reflex at admission, stage of HIE, vital sign change (respiratory rate at 24 hours, heart rate at 24 hours, temperature change at 24 hours, oxygen saturation during admission, and 24 hours, random blood sugar during admission and 24 hours), aminophylline and oxygen management were identified as a predictor for survival of asphyxiated neonates. Therefore, health professionals should be cautious and encouraged to take the proper action when a baby exhibits any vital sign change. Last but not least, researchers are urged to carry out prospective studies using current findings as a guide to having solid evidence on survival, recovery, and predictors. The ability of the medical facility to manage breathing problems, the standard of nursing care, and socioeconomic considerations are also suggested to be addressed.

### Ethical Considerations

The ethical approval was obtained from Debre Berhan University, Asrat Woldeyes Health Science Campus Institutional Review Board before its commencement. In addition, an official letter of permission was obtained from Debre Berhan Compressive Specialized Hospital before the data collection begins. The data collectors gave the interviewees an explanation of the study’s purpose, benefits, risks, discomfort, and right to withdraw or refuse at any time for any reason before the interview commenced. Before the interview began, the study participants were provided written consent. In addition, the privacy and confidentiality of participants were assured during the data collection. The individual’s identity was not revealed, so the responses to the study were anonymous.

## Data Availability

all the relevant data contained in the manuscript

## Acknowledgements

The authors would like to thank the Debre Berhan Comprehensive Specialized Hospital staff and administrators for their collaboration and unreserved help during data collection.

## Authors’ contributions

SGY: conceptualization, methodology, software, formal analysis, writing original draft, Manuscript writing

ET: supervised the design of the study, validation, data curation, writing, and editing critically

TMD: supervised the design of the study, validation, data curation, writing, and editing critically

ZAG: participate in software, data extraction, and analysis. Finally, all authors approved the manuscript.

## Declaration of conflicting interest

Regarding the research, writing, and/or publication of this article, the author(s) have declared that they have no potential conflicts of interest.

## Funding

No funding was used for this study.

## Data Sharing

Upon a reasonable request, the corresponding author will make the data utilized and/or analyzed for this study available.

---

**Figure 1:**
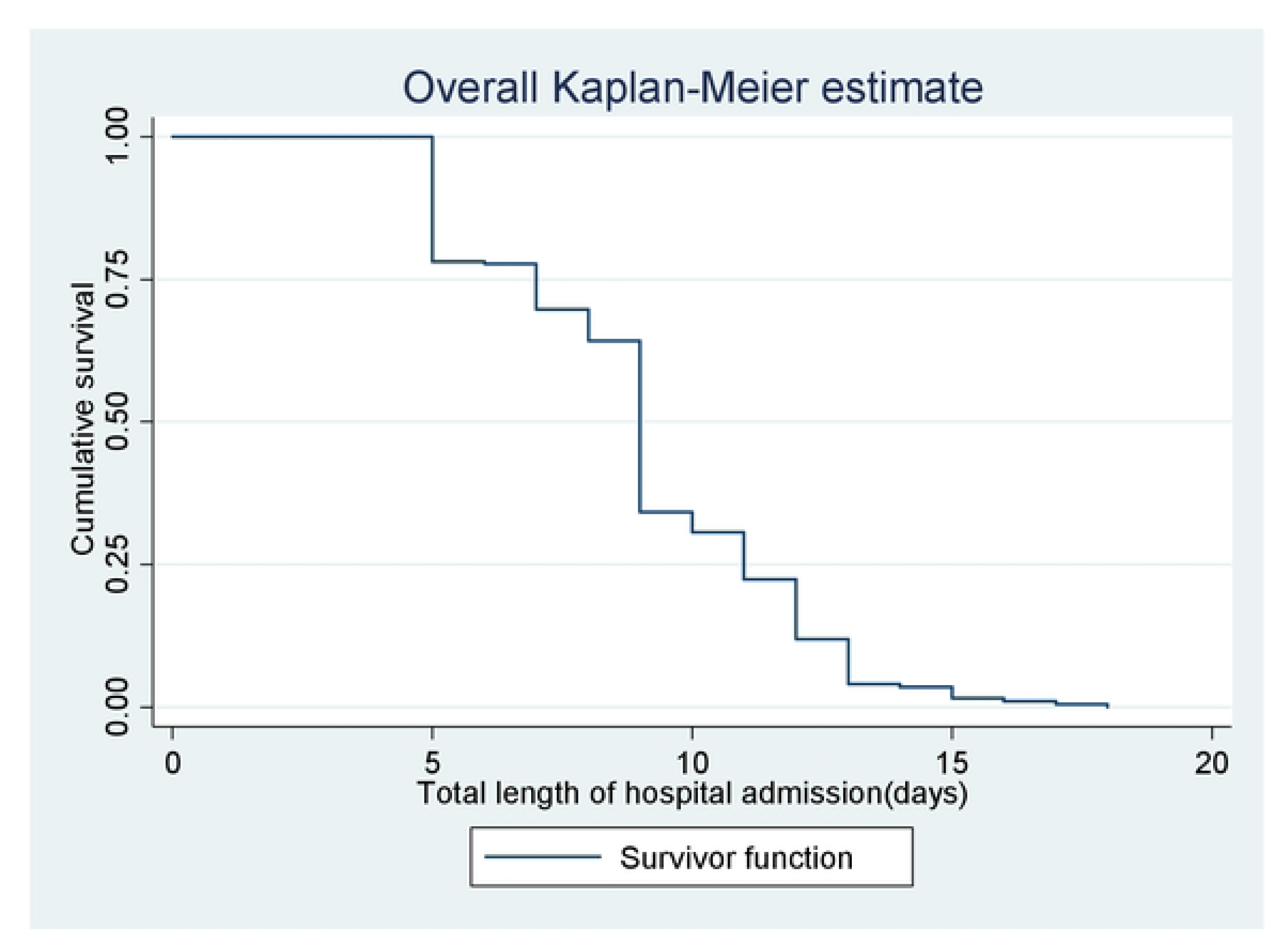
Overall Kaplan-Meier survival estimate of asphyxiated neonates admitted in DBCSH, Ethiopia from 2020-2022, 2023.

## Notes

### Competing Interest Statement

The authors have declared no competing interest.

### Funding Statement

The author(s) received no specific funding for this work.

